# Mismatch repair gene specifications to the ACMG/AMP classification criteria: Consensus recommendations from the InSiGHT ClinGen Hereditary Colorectal Cancer / Polyposis Variant Curation Expert Panel

**DOI:** 10.1101/2024.05.13.24307108

**Authors:** John Paul Plazzer, Finlay Macrae, Xiaoyu Yin, Bryony A. Thompson, Susan M Farrington, Lauren Currie, Kristina Lagerstedt-Robinson, Jane Hübertz Frederiksen, Thomas van Overeem Hansen, Lise Graversen, Ian M. Frayling, Kiwamu Akagi, Gou Yamamoto, Fahd Al-Mulla, Matthew J. Ferber, Alexandra Martins, Maurizio Genuardi, Maija Kohonen-Corish, Stéphanie Baert-Desurmont, Amanda B. Spurdle, Gabriel Capellá, Marta Pineda, Michael O. Woods, Lene Juel Rasmussen, Christopher D. Heinen, Rodney J. Scott, Carli M. Tops, Marc S. Greenblatt, Mev Dominguez-Valentin, Elisabet Ognedal, Ester Borras, Suet Y. Leung, Khalid Mahmood, Elke Holinski-Feder, Andreas Laner, the InSiGHT - ClinGen Hereditary Colon Cancer / Polyposis Variant Curation Expert Panel

**Affiliations:** Department of Colorectal Medicine and Genetics, Royal Melbourne Hospital, Parkville, Australia; Department of Medicine, University of Melbourne, Royal Melbourne Hospital, Parkville Australia; Department of Pathology, Royal Melbourne Hospital, Parkville, Australia; Colorectal Cancer Genetics Group, CRUK Edinburgh Centre, Institute of Genetics and Cancer, University of Edinburgh, Scotland; GeneDx LLC, Gaithersburg, MD, USA; Department of Clinical Genetics, Karolinska University Hospital, Stockholm, Sweden; Department of Clinical Genetics, Rigshospitalet, Copenhagen University Hospital, Copenhagen, Denmark; Department of Clinical Medicine, Faculty of Health and Medical Sciences, University of Copenhagen, Denmark; Department of Clinical Genetics, Aarhus University Hospital, Aarhus, Denmark; Inherited Tumour Syndromes Research Group, Institute of Medical Genetics, Cardiff University, CF14 4XN, UK; Department of Molecular Diagnosis and Cancer Prevention, Saitama Cancer Center, Saitama, Japan; Department of Genetics and Bioinformatics, Dasman Diabetes Institute, Kuwait; Department of Laboratory Medicine and Pathology, Mayo Clinic, Rochester, Minnesota, USA; Univ Rouen Normandie, Inserm U1245, F-76000 Rouen, France; Dipartimento di Scienze della Vita e Sanità Pubblica, Università Cattolica del Sacro Cuore, Rome, Italy; UOC Genetica Medica, Fondazione Policlinico Universitario A. Gemelli IRCCS, Rome, Italy; Woolcock Institute of Medical Research, Macquarie University, Sydney, Australia; Univ Rouen Normandie, Inserm U1245 and CHU Rouen, Department of Genetics, F-76000 Rouen, France; Population Health Program, QIMR Berghofer Medical Research Institute, Herston, Australia; Hereditary Cancer Program, Catalan Institute of Oncology, Institut d’Investigació Biomèdica de Bellvitge (IDIBELL), ONCOBELL Program, L’Hospitalet de Llobregat, Barcelona, Spain; Centro de Investigación Biomédica en Red de Cáncer (CIBERONC), Spain; Discipline of Genetics, Memorial University of Newfoundland, St John’s, NL, Canada; Center for Healthy Aging, University of Copenhagen, Copenhagen, DK-2200 Copenhagen, Denmark; Department of Cellular and Molecular Medicine, University of Copenhagen, Copenhagen, DK-2200 Copenhagen, Denmark; Center for Molecular Oncology, UConn Health, Farmington, CT, USA; Division of Molecular Medicine, NSW Health Pathology North – Newcastle, NSW 2305, Australia; The Hunter Medical Research Institute and the University of Newcastle, Newcastle, NSW, Australia; Department of Clinical Genetics, Leiden University Medical Center, The Netherlands; Department of Medicine, University of Vermont Cancer Center, Larner College of Medicine, Burlington VT, USA; Department of Tumor Biology, Institute for Cancer Research, Oslo University Hospital, Oslo, Norway; Western Norway Familial Cancer Center, Department of Medical Genetics, Haukeland University Hospital, Bergen, Norway; Invitae Corporation, San Francisco, California, USA; Department of Pathology, School of Clinical Medicine, The University of Hong Kong, Queen Mary Hospital, Pokfulam, Hong Kong SAR, China; Colorectal Oncogenomics Group, Department of Clinical Pathology, Victorian Comprehensive Cancer Centre, The University of Melbourne, Parkville, Victoria, 3010, Australia; University of Melbourne Centre for Cancer Research, Victorian Comprehensive Cancer Centre, Parkville, Victoria 3010 Australia; Melbourne Bioinformatics, The University of Melbourne, Melbourne, Victoria, Australia; MGZ – Medizinisch Genetisches Zentrum, Bayerstr. 3-5, 80035 Munich, Germany; Medizinische Klinik und Poliklinik IV, Campus Innenstadt, Klinikum Der Universität München, 80336 Munich, Germany

## Abstract

**Background:** It is known that gene- and disease-specific evidence domains can potentially improve the capability of the ACMG/AMP classification criteria to categorize pathogenicity for variants. We aimed to include gene–disease-specific clinical, predictive, and functional domain specifications to the ACMG/AMP criteria with respect to MMR genes.

**Methods:** Starting with the original criteria (InSiGHT criteria) developed by the InSiGHT Variant Interpretation Committee, we systematically addressed specifications to the ACMG/AMP criteria to enable more comprehensive pathogenicity assessment within the ClinGen VCEP framework, resulting in an MMR gene-specific ACMG/AMP criteria.

**Results:** A total of 19 criteria were specified, 9 were considered not applicable and there were 35 variations of strength of the evidence. A pilot set of 48 variants was tested using the new MMR gene-specific ACMG/AMP criteria. Most variants remained unaltered, as compared to the previous InSiGHT criteria; however, an additional four variants of uncertain significance were reclassified to P/LP or LB by the MMR gene-specific ACMG/AMP criteria framework.

**Conclusion:** The MMR gene-specific ACMG/AMP criteria have proven feasible for implementation, are consistent with the original InSiGHT criteria, and enable additional combinations of evidence for variant classification. This study provides a strong foundation for implementing gene–disease-specific knowledge and experience, and could also hold immense potential in a clinical setting.

## 1 Introduction

Lynch syndrome, previously referred to as hereditary nonpolyposis colorectal cancer, is caused by a heterozygous constitutional pathogenic loss of function of a mismatch repair (MMR) gene, which is either *MLH1* (OMIM 120436), *MSH2* (OMIM 609309), *MSH6* (OMIM 600678), or *PMS2* (OMIM 600259).

It can also arise from the deletion of the 3’end of *EPCAM* (TACSTD1), which results in hypermethylation of the *MSH2* promoter. These factors foster the accumulation of somatic mutations that cannot be corrected by the defective machinery, resulting in tumors with a microsatellite instability (MSI)-high phenotype. Although the tumor spectrum is variegated, colorectal or endometrial cancers predominate. Strategies aimed at identifying carriers of pathogenic variants range from ascertainment of family history, testing for MMR-deficient cancers in patients, to universal constitutional sequencing of patients presenting with colorectal (Tiwari, 2016) or endometrial cancers (Kahn, 2019).

A public database of germline MMR variants and variants involved in other familial gastrointestinal (GI) cancer syndromes has long been maintained by the International Society for Gastrointestinal Hereditary Tumors (InSiGHT), the preeminent professional body representing healthcare workers, researchers, and medical geneticists interested in familial GI cancer. The InSiGHT databases for inherited GI cancer syndromes centralize data and aid expert variant classification (Plazzer, 2013). Since the 1990s, MMR variants have steadily accrued, with changes in technology accompanying significant increases in the identified variants. By 2023, the number of unique constitutional MMR variants in public variant databases reached over 6,500 in LOVD and over 23,000 in ClinVar. In 2014, the InSiGHT Variant Interpretation Committee criteria for the classification of MMR variants were published (Thompson, 2014), and the committee was thereafter recognized as an external expert panel by ClinGen (Rehm, 2015). The landmark American College of Medical Genetics and Genomics/Association for Molecular Pathology (ACMG/AMP) variant classification criteria were published in 2015 (Richards, 2015), proposing generic classification criteria requiring gene or disease specifications. They were soon adopted by many gene–disease expert groups, including those within the Variant Curation Expert Panel (VCEP) framework developed by ClinGen (Rivera-Muñoz, 2018). Latterly these have been recognized by the Food and Drug Administration (FDA) as a regulated source for curating variants.

Following an agreement between InSiGHT and ClinGen, the Hereditary Colorectal Cancer/Polyposis VCEP (https://clinicalgenome.org/affiliation/50099/) was formed out of the existing InSiGHT Variant Interpretation Committee, and was expanded to other genes predisposing hereditary GI cancer, including *APC, MUTYH, STK11, POLD1*, *POLE, SMAD4,* and *BMPR1A*, for which there is substantial expertise within the membership of InSiGHT. As this VCEP covers multiple genes for several hereditary GI tumors, it has been divided into sub-VCEP groups for organizational efficiency. Here, we describe the work of the MMR VCEP subgroup in adapting gene-specific ACMG/AMP criteria and the validation of the criteria through pilot variant classification.

## 2 Methods

### 2.1 InSiGHT-ClinGen MMR VCEP

We made an application to be recognized as a ClinGen internal VCEP to ClinGen, which was approved by ClinGen in February 2021. This involved listing of the group’s members, expertise, and scope of our curation activity. Members of the InSiGHT ClinGen Hereditary Colorectal Cancer/Polyposis Variant Curation Expert Panel (https://clinicalgenome.org/affiliation/50099/) have diverse technical and clinical expertise from the private, public, research, and healthcare sectors. There is a broad international representation of GI surgeons, genetic counsellors, medical geneticists, oncologists, scientists, bioinformaticians, and clinical laboratory diagnosticians.

### 2.2 Gene–disease specifications to the ACMG/AMP criteria

Previously, the InSiGHT criteria (https://www.insight-group.org/criteria/) used predetermined combinations of evidence and a Bayesian analysis component, which incorporated *in silico* prediction, tumor characteristics, functional assay, and variant-disease segregation (Thompson, 2013). These criteria inform the new ACMG/AMP specifications for MMR genes. The ACMG/AMP framework uses weight of evidence corresponding to Supporting, Moderate, Strong, and Very Strong categories (Figure 1). Furthermore, these categories are calibrated against likelihood ratio (LR) thresholds from modelling that align the ACMG/AMP criteria with a Bayesian probability framework (Tavtigian, 2018). Therefore, while both current and original InSiGHT criteria utilize Bayesian methods, the new criteria integrate the evidence combinations and Bayesian probabilities into a single framework. Our ACMG/AMP specifications are updated periodically; to find the latest information, please visit: https://cspec.genome.network.

**Figure 1.**
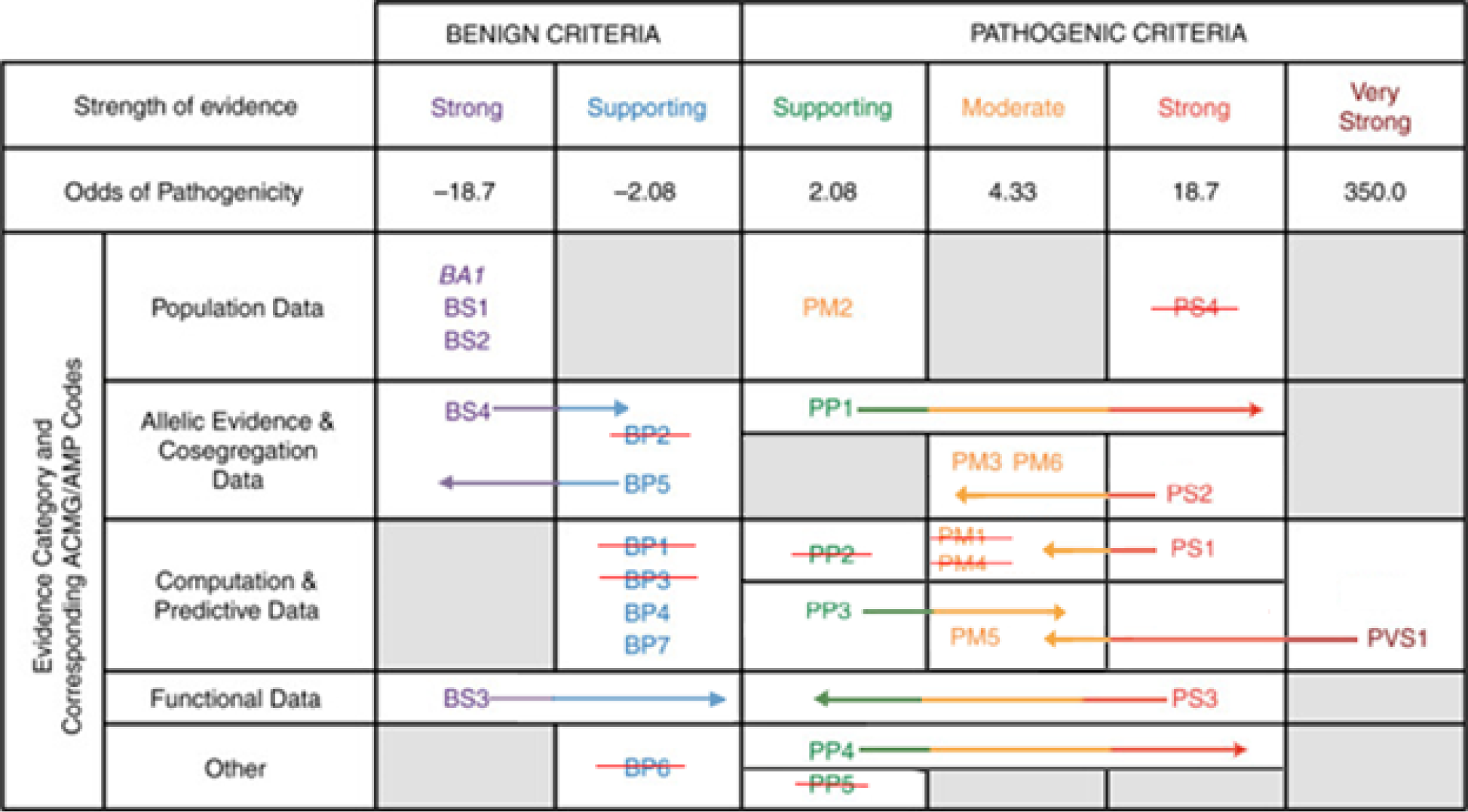
Specifications of the ACMG/AMP criteria. Arrows indicate the range of strength of the evidence, adapted from (Strande, 2018). BA1 is included with Strong criteria even though it is Stand Alone strength. PM3, PM6, and PS2 are subject to strength changes using a points-based system.

### 2.3 LS penetrance

The ACMG/AMP criteria can classify high-penetrance MMR variants for LS, while comparatively low- or moderate-risk variants cannot be classified owing to scarcity of data (Dominguez-Valentin, 2021). *PMS2* variants are an exception, as this gene is recognized for having low-penetrance; therefore, loss of function variants are presumed to confer a low risk of cancer in carriers of heterozygous *PMS2* pathogenic variants. Information on risks of cancer according to organ, age, gender, and gene (Dominguez-Valentin, 2020) is available on the Prospective Lynch Syndrome Database (PLSD: www.PLSD.eu). Estimates of the penetrance of LS also inform segregation analysis by accounting for LS-specific cancer risk, dependent on the age of the patient at diagnosis and their sex. This VCEP recommends the COsegregation OnLine v3 tool (Belman, 2020), which incorporates penetrance for MMR genes and has a free public web interface (https://fengbj-laboratory.org/cool2/manual.html).

### 2.4 Inclusion of tumor characteristics

One of the hallmarks of LS, observed in respective tumors, is the microsatellite instability (MSI) and MMR immunohistochemistry (IHC) pattern demonstrating a loss of protein expression. Loss of MMR protein expression is highly concordant (98%) with the MSI status in LS, though MSI more commonly occurs outside the context of LS in sporadic colorectal or endometrial cancer (Latham, 2019). It is also recognized that individuals with LS can still develop sporadic microsatellite stable (MSS) cancers, which is not due to their constitutional variant. In a study (Li, 2020) on MSI/IHC status and the application of variant classification, the researchers achieved 98% accuracy in variant classification when combined with *in silico*, clinical/family history, co-segregation, allele frequency, and co-occurrence data. The ACMG/AMP criteria incorporates these data types and the updated tumor characteristic likelihood ratios from the study by Li *et al*. MSI/MSS and IHC data are specified in the criteria for colorectal and endometrial cancers owing to existing calibration studies of these particular tumor characteristics with germline pathogenic variants. Reciprocally, tumor IHC is also specified for any LS spectrum tumor having protein expression that is inconsistent with the gene demonstrating the variant.

### 2.5 Functional assay specifications

New recommendations for the application of the functional evidence types PS3/BS3 (Brnich, 2020) assign the strength of the ACMG/AMP evidence, depending on the level of statistical analysis used for assay validation. As per the Brnich *et al*. recommendation, at least 11 controls are required to reach a Moderate level evidence and a rigorous statistical analysis to reach a Strong level of evidence. There are currently three assays recognized with calibrated odds of pathogenicity that reach a Strong level of evidence (PS3), namely, the Complete *in vitro* MMR Assay (CIMRA) for *MLH1*, *MSH2*, *MSH6*, and *PMS2* (Drost, 2019), (Drost, 2020), (Rayner, 2022), a deep mutational scan of MSH2 (Scott, 2022) (Jia, 2021), and a cell-based assay for *MLH1* (Rath A, 2022). These assays employed rigorous statistical analysis calibrating the level of *in vitro* MMR activity to the corresponding probability of pathogenicity. They have, therefore, been validated to reach Supporting, Moderate, or Strong level of evidence for PS3 and Supporting or Strong for BS3 evidence (Supplementary Table 1).

In addition to the calibrated functional assays, a decision tree (Figure 2) logically combines other types of functional assay to test different aspects of MMR function, such as protein expression, subcellular localization, and RNA splicing, to determine if a variant leads to proficient or deficient function. It was published in 2014 (Thompson, 2014) and updated in 2020 (Thompson, 2020), and is now incorporated into the VCEP MMR criteria, though calibrated assays are recommended for prospective variant analysis. For the assays detailed in the functional assay decision tree, as per Brnich *et al,* the criterion of a minimum number of 11 pathogenic and benign controls has been met for each type of assay (Supplementary Table 1); this excludes splicing assays, which have specific requirements in the criteria descriptions. The decision tree is used for interpreting previous or historical assay results, not as a guide for performing prospective assays of which we recommend the calibrated assays.

**Figure 2.**
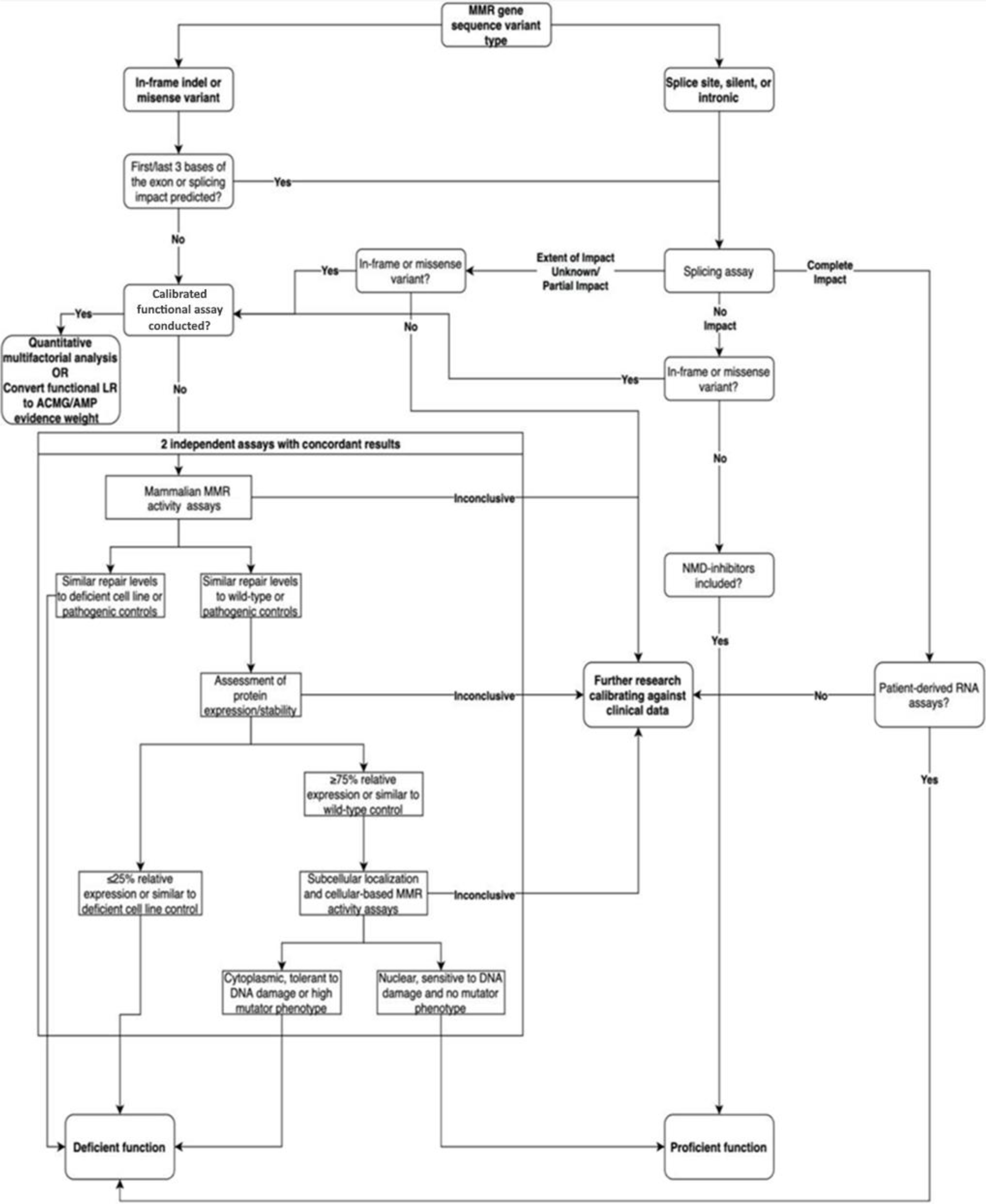
Flowchart demonstrating the interpretation of functional assay data, adapted from Thompson et al. 2020. If RNA splicing is used for PVS1, it should not be used for PS3.

### 2.6 In silico variant effect predictions tools

*In silico* meta-predictor tools and tools calibrated for all missense variants in specific genes offer improved performance, as compared to individual predictors (Tian, 2019) (Pejaver, 2022). Here, the PP3/BP4 criteria for *in silico* evidence were derived from previous analyses described in (Thompson, 2013). Briefly, Thompson et.al. (2013) calibrated individual *in silico* variant effect prediction tools against MMR variants with designated pathogenicity, showing that a combination of MAPP-MMR and Polyphen-2 predictors showed higher performance, in terms of specificity and sensitivity, as compared to other individual tools. For MLH1, MSH2, and MSH6 missense variants, pre-computed prior probabilities are available online (https://hci-priors.hci.utah.edu/PRIORS/) that map to Supporting, Moderate, or Strong evidence. Similarly, for PMS2 missense variants, we recommend using the HCI “MAPP/PP2 Prior P” predictions available at http://hci-lovd.hci.utah.edu/variants.php?select_db=PMS2_priors. For intronic and synonymous variants, we recommend predicting splice altering consequence with SpliceAI (Riepe, 2021).

### 2.7 Pilot variant classification

A series of teleconferences between 2020 and 2021 were held to merge the existing InSiGHT MMR-specific criteria with the ACMG/AMP criteria in a ClinGen VCEP framework. Feedback from the ClinGen Sequence Variant Interpretation Working Group (SVI WG) was incorporated into the criteria, and the SVI WG approved the criteria in May 2022. Following the creation of the initial draft of the criteria, a pilot batch of 48 variants were manually selected from the InSiGHT and ClinVar databases to assess whether their classification could be changed from variant of uncertain significance (VUS), likely benign (LB), or likely pathogenic (LP), using the new criteria and/or new information. The variants were curated for this study using existing information from public databases and literature, the Colon Cancer Family Registry (www.coloncfr.org), as well as information from VCEP members which are available on LOVD. The types of variants included intronic, splice site, missense, synonymous, in-frame deletion, nonsense, and frameshift variants. Variants were reviewed independently by experts and biocurators, followed by VCEP teleconference discussions. Disparities between reviewers were discussed and appropriate revisions to the criteria were made. Biocurators entered the variants, the ACMG/AMP criteria that was met, summary text and classifications into the ClinGen Variant Curation Interface tool (Preston, 2022) to obtain a 3-star VCEP classification status within the FDA-recognized ClinGen framework.

### 2.8 Selection of transcripts

The preferred MANE Select transcripts are NM_000249.4 (*MLH1*), NM_000251.3 (*MSH2*), NM_000179.3 (*MSH6*), and NM_000535.7 (*PMS2*). Variants are described using the Human Genome Variation Society (HGVS) nomenclature (https://varnomen.hgvs.org/).

## 3 RESULTS

The MMR-specific ACMG/AMP criteria are listed on https://cspec.genome.network. A total of 19 criteria are specified, with 35 variations of the criteria when including the different strength options. Of these, nine were not used (PS4, PM1, PM4, PP2, PP5, BP1, BP2, BP3, and BP6). The criteria were the same for all four MMR genes, unless indicated otherwise. The allowable combinations of evidence were also updated based on the ClinGen Sequence Variant Interpretation Recommendation for PM2, Version 1.0 (https://clinicalgenome.org/working-groups/sequence-variant-interpretation/) and the findings from Tavtigian et al.

### 3.1 Variant Allele Frequency thresholds (PM2, BA1, and BS1)

The population allele frequency of a variant represents the frequency or incidence of the variant in a reference population, such as the genome aggregation database (gnomAD) (Karczewski, 2020). The ACMG/AMP criteria are met when the frequency of a population allele is higher than expected for disease-causing variants (BA1 and BS1), or reciprocally, are rare or non-existent in the reference population, which can imply a possible pathogenic role of the variant (PM2). The CardioDB allele frequency calculator allows the calculation of allele frequency thresholds for both Benign Stand Alone (BA1) or Strong (BS1) evidence for a disease of interest, with appropriate input parameters (Whiffin, 2017). These thresholds were adapted from a study that calculated allele frequency thresholds at 99% confidence for each MMR gene (Canson, 2022), using published penetrance and prevalence values. For pathogenic PM2_supporting evidence to be met, the threshold is set at 1/50,000 alleles (Grpmax filtering allele frequency of 0.00002 in gnomAD v4.1.0 dataset).

### 3.2 Variant type and location (PVS1, PS1, and PM5)

We modified the PVS1 flowchart using recommendations from Tayoun et al. (Tayoun, 2018) to reflect the current understanding of pathogenesis from loss of function variants within MMR genes. We created a simplified version of the PVS1 decision tree (Figure 3) and show the logic for assigning the ACMG/AMP criteria to truncating, canonical splice site (intervening sequence (IVS)±1/2 invariant nucleotides within an intron), confirmed splice defect, large genomic alteration, and initiation codon variants.

**Figure 3.**
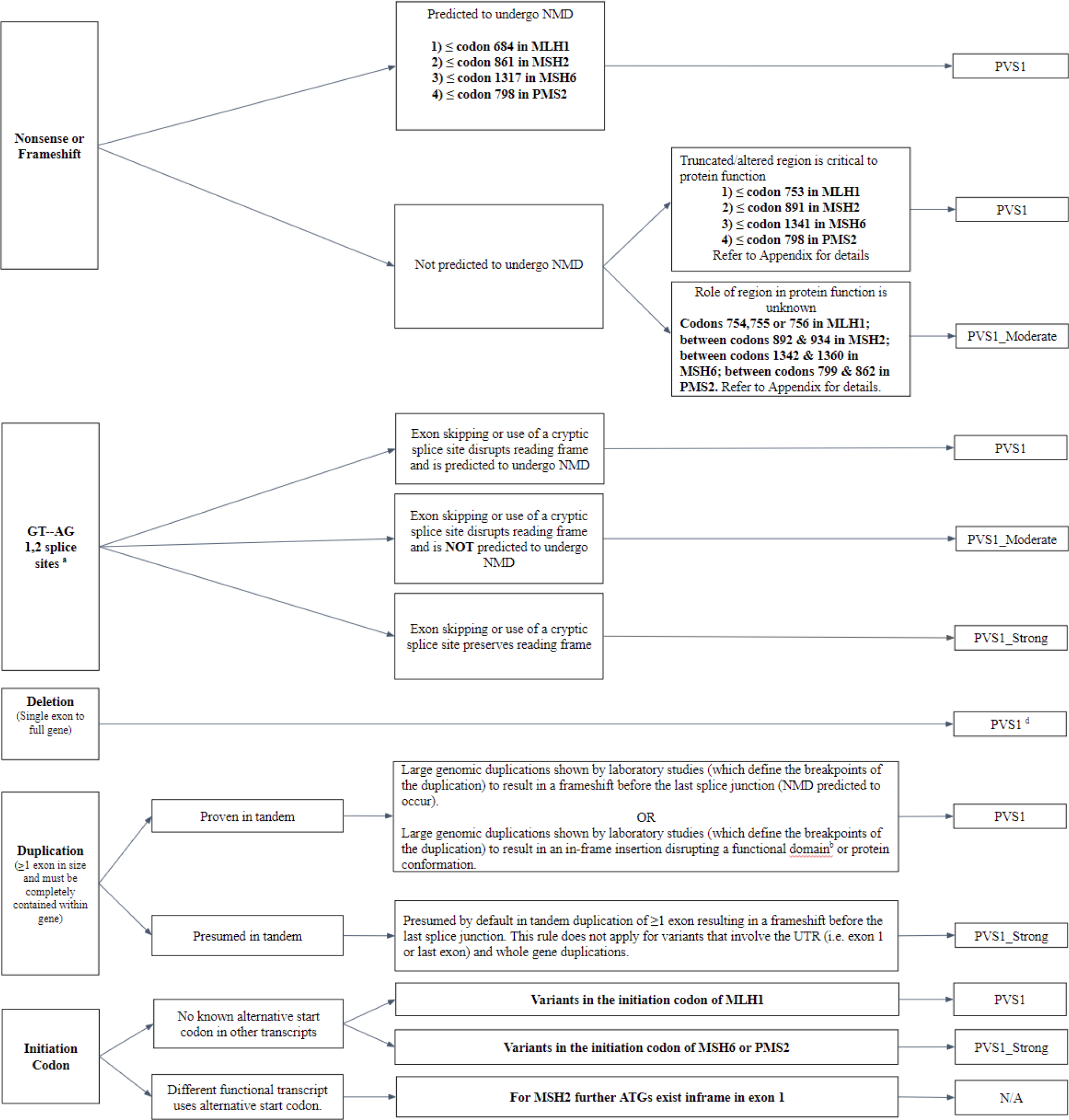
The PVS1 decision tree, adapted from Tayoun et al. 2018.

#### 3.2.1 Nonsense/frameshift variants

PVS1 is assigned if the variant is predicted to encode a protein termination codon located at least 50 bp before the last exon–exon junction, as per the nonsense-mediated mRNA decay (NMD) rule (Lewis, 2003), or, if available, before the reference codon of the most distal (3’) truncating variant, unambiguously classified as pathogenic in the last exon of the gene of interest (Figure 3). For MMR genes, the truncated/altered region that is critical to protein function is defined as ≤ codon 753 in *MLH1* using the location of known pathogenic variant *MLH1*: c.2252_2253del; ≤ codon 891 in *MSH2* using the location of known pathogenic variant *MSH2*: c.2662del; ≤ codon 1341 in *MSH6* using the location of known pathogenic variant *MSH6*: c.3984_3987dup; and ≤ codon 798 in *PMS2* using the ≥50 nucleotide NMD-rule.

**Figure 3.**
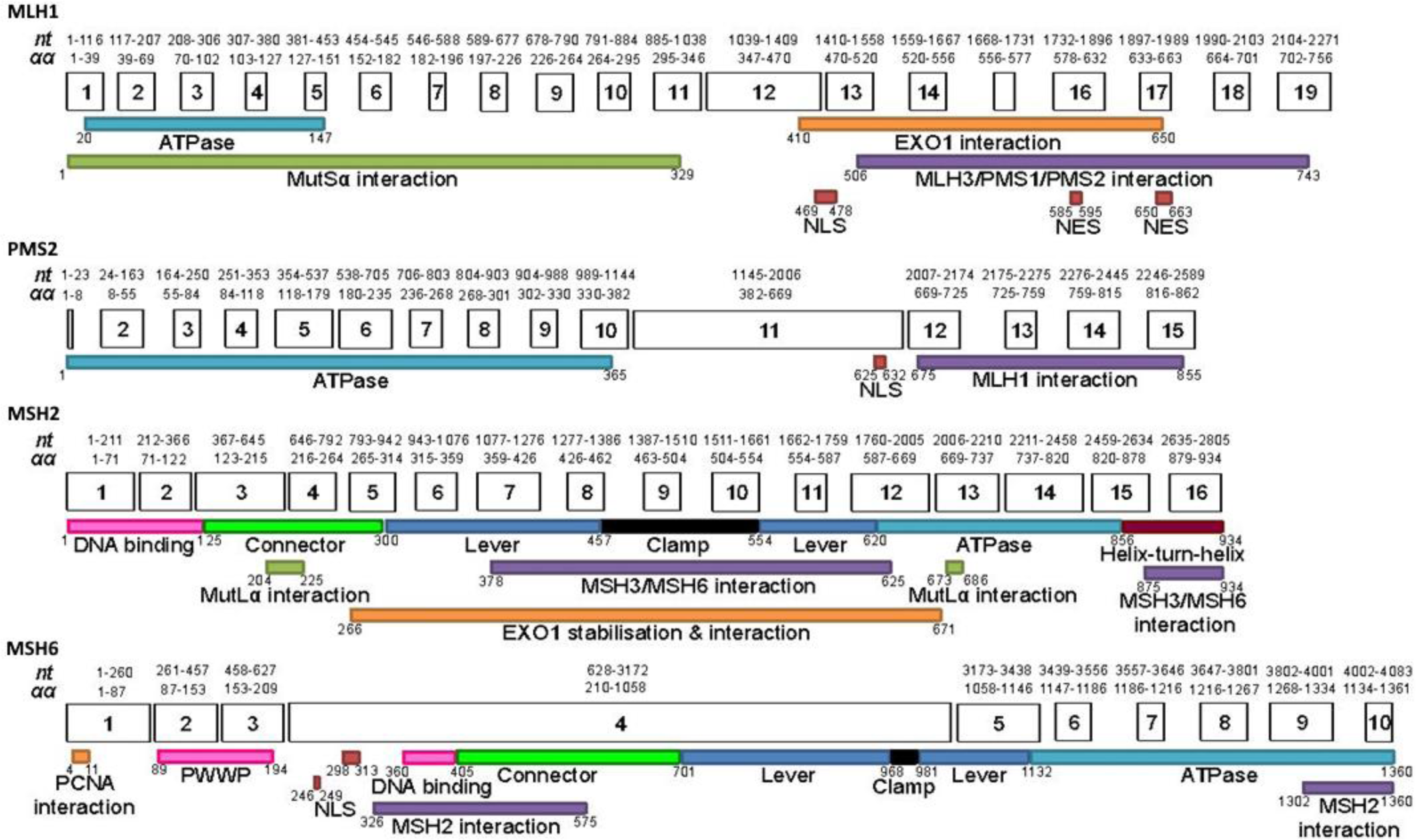
Linear schematic of mismatch repair gene functional domains according to amino acid positions. Adapted from InSiGHT criteria v2.4 (https://www.insight-group.org/content/uploads/2018/08/2018-06_InSiGHT_VIC_v2.4.pdf)

#### 3.2.2 Splicing aberrations

Variants affecting the canonical splice sites (IVS±1/2) were previously considered pathogenic due to their likely impact on splicing. The ACMG/AMP recommendations by Tayoun et al. provide additional nuance and fine-tuning of the strength of PVS1 based on whether the impact is in-frame, out-of-frame, and/or NMD-based. For canonical splice site variants, PVS1 is met if the predicted splicing alteration results in a frameshift mutation and NMD is predicted. If exon skipping or cryptic splice site usage preserves the reading frame, then PVS1_Strong is to be applied. If exon skipping or cryptic splice site usage disrupts the reading frame, and is not predicted to undergo NMD, then PVS1_Moderate is applied, as per the guidelines of Tayoun et al. PP3 cannot be used together with this criterion as the variant type already incorporates Very Strong prediction. For splicing aberrations confirmed using patient mRNA, PVS1 can be assigned where additional requirements are met, including complete defect (no full-length transcript produced from the variant allele) and confirmation in a minigene assay or an additional RNA assay from an independent laboratory, if it is not a predicted splice site variant.

#### 3.2.3 Large genomic rearrangements

MMR exon deletions are considered to meet PVS1 due to deletion of functional domains, altered reading frame, and/or truncation leading to NMD. Exon duplications are presumed to be in tandem (Richardson, 2019) and, if predicted to result in a frameshift with NMD, are considered as PVS1_Strong, as described in Tayoun et al. Whole gene duplications and other duplications affecting the first and last exons are excluded from this criterion.

#### 3.2.4 Variants in the initiation codon

Variants in the initiation codon of *MLH1* are deemed to meet PVS1 owing to previously known and well established pathogenic variants at this location (Zhang, 2018). For *MSH6* and *PMS2* initiation codons, PVS1_Strong is recommended due to pathogenic variants reported on LOVD (Fokkema, 2021). For *MSH2*, alternative ATGs exist inframe in exon 1 (Cyr, 2012), so this criterion is not applicable at any evidence weight.

#### 3.2.5 Substitution at same position as a previously classified pathogenic variant (PS1, PM5)

PS1 and PM5 criteria seek to assign evidence based on the pathogenic classification of another variant at the same location. PS1 is Strong evidence derived from another pathogenic variant that causes the same presumed amino acid change or splice site impact. For missense variants, it is required that a splicing defect is neither predicted nor the basis of the other variant’s classification. The same presumed amino acid change for both variants implies the same impact on protein function. PM5 is similar except the previously classified variant is for a different amino acid at the same position, with an additional caveat that *in silico* PP3 is met and only at a Supporting level. This is to ensure the variant to be classified is predicted to be damaging while avoiding overuse of *in silico* predictions at this location if PP3_Moderate was met. For variants affecting the same non-canonical splice site nucleotide as a confirmed pathogenic splice-site variant with similar or worse splicing *in silico* predictions, PS1 is specified.

### 3.3 In silico predictions (PP3, BP4, AND BP7)

Computed predictions based on combined MAPP-MMR and MaxEntScan predictions for all MLH1, MSH2, and MSH6 missense variants are available online at https://hci-priors.hci.utah.edu/PRIORS/ and therefore can be converted to ACMG criteria evidence using the Bayesian framework LR thresholds. For synonymous and intronic variants, the MMR VCEP approves the use of SpliceAI to predict the impact on splicing. Synonymous and deep intronic variants (past +7/-21 nucleotides within an intron) with predicted non-impact on splicing can be classified as LB using BP4 and BP7.

### 3.4 Clinical data-driven rules

#### 3.4.1 Phenotype and segregation data (PP4, PS2, PM3, PM6, PP1, BP5, BS2, and BS4)

Phenotype evidence is based on the MSI/MMR IHC pattern in colorectal or endometrial cancer tumors, which are a hallmark of LS. The number of tumors required correspond to the odds calculated in published methods (Thompson, 2013) (Thompson, 2014) and have been recently updated (Li, 2020). Briefly, MSI and somatic *BRAF* mutation characteristics were used to assign odds ratios to tumor phenotypes, derived as the ratio of characteristics of known pathogenic carrier cases *versus* that of known non-carrier cases. Finally, the number of tumors required to meet the odds ratios, as defined in the Bayesian framework of the ACMG/AMP criteria, were assigned the corresponding strength of the ACMG/AMP criteria.

For PP4 evidence, the number of colorectal or endometrial cancer tumors is counted; whereby, one MSI-H tumor is PP4 (Supporting), two is Moderate, and three or more is Strong. Multiple tumors can be included from an individual patient if they are independent tumors. Somatic MLH1 promoter methylation should be excluded in MLH1-/PMS2-tumors. Loss of expression of MMR genes by IHC evidence should be consistent with the variant gene and the protein that is tested and must take into account the MutSα and MutLα heterodimers: MLH1 and PMS2 loss is consistent with an *MLH1* pathogenic variant, MSH2 and MSH6 loss is consistent with an *MSH2* pathogenic variant, MSH6 loss is consistent with an *MSH6* pathogenic variant, and PMS2 loss is consistent with a *PMS2* pathogenic variant.

BP5 requires two colorectal or endometrial cancer tumors with MSS and/or no loss of MMR protein expression, or LS-spectrum tumors with a loss of MMR protein(s) thatis inconsistent with the variant gene. BP5 can also be met with one *BRAF* V600E mutation tumor (colorectal cancer alone) or MLH1 methylation (in LS-spectrum tumor alone) with MSI-H/MLH1 loss. For BP5, MSS is an indicator of a benign role for the variant. Originally, only one MSS tumor was required to meet BP5, as per the odds ratio calculations. However, upon classifying several variants as LB with one tumor combined with *in silico* BP4, the VCEP decided that this could lead to unwarranted LB classification. Thus, while the Bayesian analysis may indicate a benign classification, the experience of clinical experts overruled quantitatively derived evidence in this instance. This is in contrast to the PP4 criteria, which does allow a single tumor to meet the criteria. This may be attributed to the fact that benign criteria are fewer in number and are restricted in strength (Supporting or Strong only); therefore, more stringent criteria are required to meet a benign classification. It may also reflect the possibility that a single incorrect clinical result for tumor characteristics, due to technical issues, phenocopies, or errors, could significantly alter the original Bayesian likelihood analysis. To resolve this issue, it was agreed that two MSS tumors are required to meet BP5 and four are required for BP5_Strong.

#### 3.4.2 Co-segregation odds to assign PP1 (Supporting, Moderate, and Strong) and BS4 (Supporting and Strong)

Co-segregation odds ratios were used previously for the InSiGHT criteria and are incorporated into the ACMG/AMP criteria. Recently, a web-based tool COsegregation OnLine v3 (<u>COOL v3 Manual</u>), enables a relatively easy way of calculating the co-segregation odds for a given pedigree and, thereby, mapping this to the ACMG/AMP criteria at different strength levels (Belman, 2020). It should also be noted that multiple families with the same variant can have their co-segregation odds multiplied together to produce a more informative result, either in favor or against pathogenicity. For PP1_Strong, there is a requirement for two or more families to be met so that a pathogenic (P/LP) classification is not reached based on data from one family alone.

#### 3.4.3 De novo variants (PS2/PM6)

Criteria for *de novo* variants are based on the 2018 ClinGen Sequence Variant Interpretation Recommendation for *de novo* Criteria (PS2/PM6) guidelines, Version 1.1 (https://clinicalgenome.org/working-groups/sequence-variant-interpretation/), for a points-based scale using tumor characteristics (MSI/IHC) from any LS-spectrum tumors to meet the criteria for pathogenic evidence. This criteria allows multiple cases of *de novo* variants to be combined using the points-based system (Table 2).

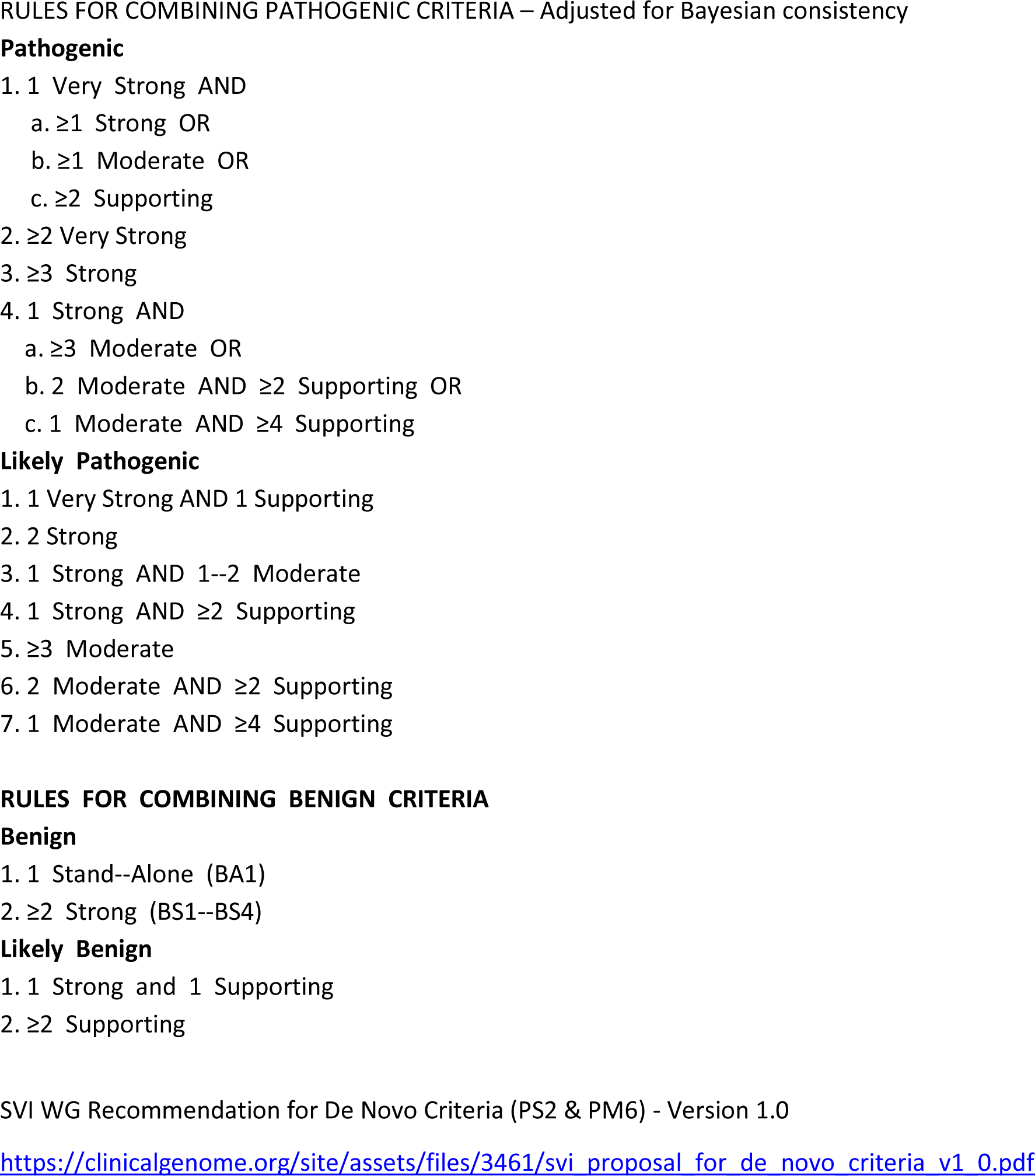

**Table 2.**
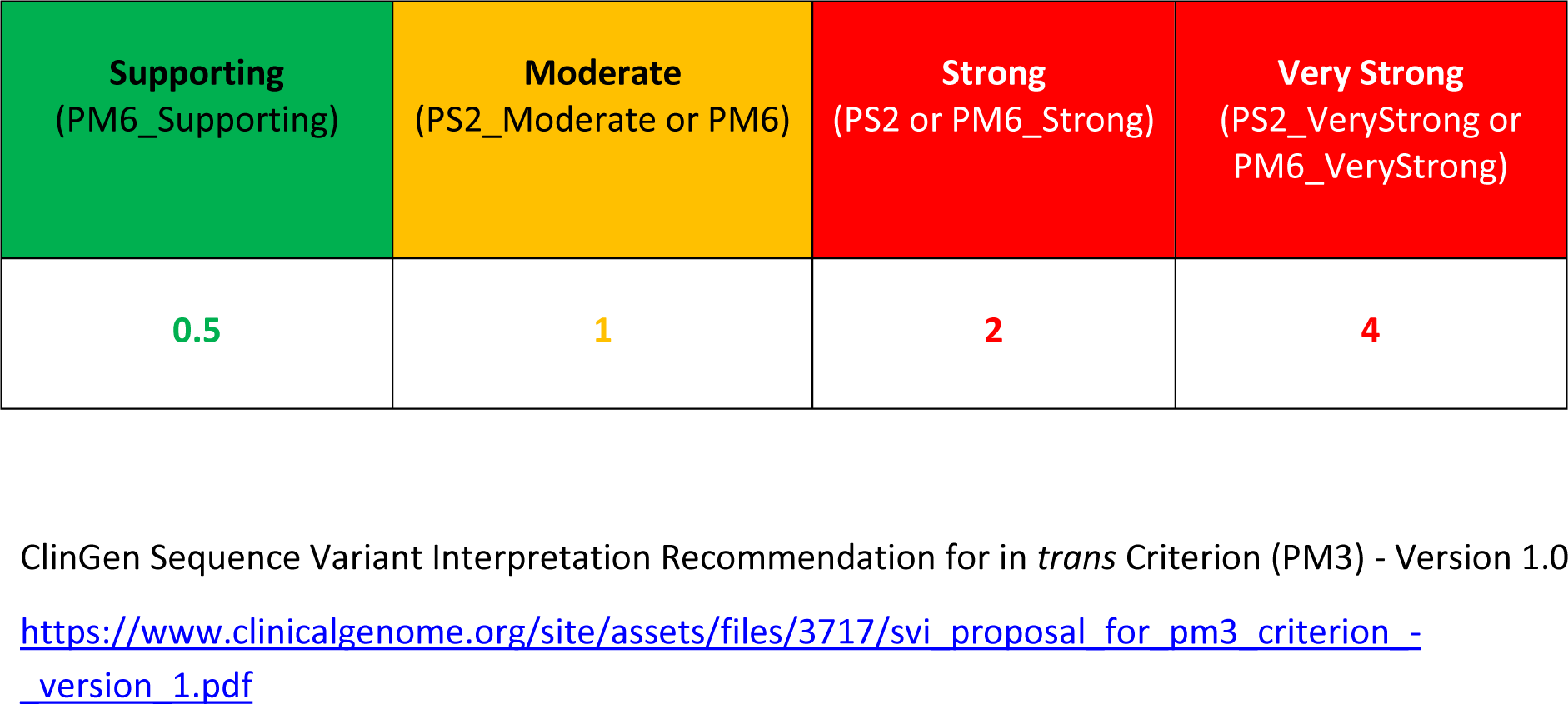
The combined point value of all *de novo* occurrences used to determine the applicable evidence strength level.

#### 3.4.4 Co-occurrence with a known P/LP variant (PM3, BS2)

For classification of variants seen in LS-suspected cases, evidence towards or against pathogenicity can be assigned (Table 3) if the variants co-occur *in trans* with known P/LP variants in the same gene. The evidence is in a benign or pathogenic direction, depending on whether cases have constitutional mismatch repair deficiency (CMMRD), which is caused by biallelic pathogenic MMR variants and predisposition to childhood cancer. A variant in cases of CMMRD can be assigned to PM3 if the CMMRD international working group recommendations are met (Table 4), as recommended by the CMMRD international working group (Aronson, 2022). Strong evidence for a benign role (BS2) is recommended if the CMMRD phenotype is not present in patients with an LS-associated cancer, over 45 years of age and with a known pathogenic variant in *trans* in the same gene. Careful consideration is required of cancer risk for MSH6 and PMS2 variants, as heterozygous variants in these genes may not significantly increase disease risk and the CMMRD phenotype may not be expressed or may be expressed at a later stage.

**Table 3.** Recommendation for determining the appropriate ACMG/AMP evidence strength level for *in trans* occurrence(s)

| **Supporting**<br>(PM3_Supporting) | **Moderate**<br>(PM3) | **Strong**<br>(PM3_Strong) | **Very Strong**<br>(PM3_VeryStrong) |
| --- | --- | --- | --- |
| **0.5** | **1** | **2** | **4** |

**Table 4:** Scoring system for aiding CMMRD diagnosis from C4CMMRD (adapted from Aronson et al 2022; PMID: 33622763)

|  |  |
| --- | --- |
| *Malignancies/premalignancies: one is mandatory; if more than one is present in the patient, add the points:* | **>=3 points = CMMRD features meets PM3 criteria after excluding the diagnosis of NF1 or LFS as individuals with those disorders could easily get to 3 points.** |
| Carcinoma from the LS spectrum* at age <25 years. | 3 points |
| Multiple bowel adenomas at age <25 years and absence of APC/MUTYH mutation(s) or a single high-grade dysplasia adenoma at age <25 years. | 3 points |
| WHO grade III or IV glioma at age <25 years. | 2 points |
| NHL of T cell lineage or sPNET at age <18 years | 2 points |
| Any malignancy at age <18 years. | 1 point |
| *Additional features: optional; if more than one of the following is present, add the points:* |  |
| Clinical sign of NF1 and/or ≥2 hyperpigmented and/or hypopigmented skin alterations Ø>1 cm. | 2 points |
| Diagnosis of LS in a first-degree or second-degree relative. | 2 points |
| Carcinoma from LS spectrum* before the age of 60 in a firstdegree, second-degree or third-degree relative. | 1 point |
| A sibling with carcinoma from the LS spectrum*, high-grade glioma, sPNET or NHL. | 2 points |
| A sibling with any type of childhood malignancy. | 1 point |
| Multiple pilomatricomas in the patient. | 2 points |
| One pilomatricoma in the patient. | 1 point |
| Agenesis of the corpus callosum or non-therapy-induced cavernoma in the patient. | 1 point |
| Consanguineous parents | 1 point |
| Deficiency/reduced levels of IgG2/4 and/or IgA. | 1 point |

\*Colorectal, endometrial, small bowel, ureter, renal pelvis, biliary tract, stomach, bladder carcinoma
CMMRD, constitutional mismatch repair deficiency; LS, Lynch syndrome; NF1, neurofibromatosis type 1; NHL, non-Hodgkin's lymphoma; sPNET, supratentorial primitive neuroectodermal tumours.

### 3.5 Experimental data-driven rules for PS3 (Supporting, Moderate, and Strong) and BS3 (Supporting and Strong)

Functional assay data that follows the functional assay flowchart (Figure 2) to arrive at a deficient or proficient function can meet PS3_Moderate or BS3_Moderate. The functional assay flowchart is a general framework for evaluating functional assays that were already performed, or from historic publications, and is not recommended for prospective studies on variants. The VCEP recommends use of the calibrated assays for prospective testing. For calibrated assays, variants can reach a maximum of PS3/BS3 level. Other assays were validated with the recommendations of Brnich et al., as per Supplementary Table 1. For missense or in-frame indel variants, MMR activity, protein expression/stability, and subcellular location are required to determine if the function is deficient or proficient, when complete splicing impact is not shown.

### 3.6 Unused criteria (PS4, PM1, PM4, PP2, PP5, BP1, BP2, BP3, and BP6)

PS4 - Due to the availability of tumor IHC data for variant classification (see PP4), PS4 has not been utilized for MMR variant classification using proband counting.

PM1 - Located in a mutational hot spot and/or critical and well-established functional domain. There are no recognized mutational hot spots that could be used for classification purposes. While there are functional domains in the MMR genes, the distribution of pathogenic variants is generalized over all the domains (unpublished data).

PM4 - Protein length change from an in-frame variant is not used due to lack of evidence.

PP2 - Missense variant in a gene with low rate of benign missense changes does not apply.

PP5 - Not recommended by ClinGen/SVI WG (Biesecker, 2018).

BP1 - Missense variant in a gene where only loss of function causes disease is not applicable.

BP2 - BS2 is used instead.

BP3 - In-frame deletions/insertions in a repetitive region without a known function is not used.

BP6 - Not recommended by ClinGen/SVI WG (Biesecker, 2018).

### 3.7 Validation through pilot variant classification

The set of 48 variants in the pilot batch covered a range of variant types commonly encountered in genetic testing, including 27 presumed missense, 3 presumed synonymous, 3 last base of exon single nucleotide variant, 8 splice site, 4 intronic, 1 in-frame deletion, 1 nonsense, and 1 frameshift (Table 5). Final consensus classifications show that 28 variants (58%) reached a P/LP classification and 12 (25%) reached a B/LB classification; the remaining eight (17 %) were VUS.

**Table 5:** Comparison of ACMG/AMP and InSiGHT criteria classifications.

| Variant | Predicted protein change | Variant type | ACMG/AMP criteria met | ACMG/AMP Class | InSiGHT Class | Concord/Disco rd |
| --- | --- | --- | --- | --- | --- | --- |
| MLH1:c.42A>C | p.(Thr14=) | Synonymous | BS1, BP4, BP7, BS4 | Benign | Benign | Concordant |
| MLH1:c.1732-19T>A |  | Intronic | BA1, BS3 | Benign | Likely Benign | Concordant |
| MSH6:c.107C>T | p.(Ala36Val) | Missense | BS1, BP4, BS2 | Benign | Likely Benign | Concordant |
| MLH1:c.649C>T | p.(Arg217Cys) | Missense | BS3, BS1, BP5 | Benign | Likely Benign | Concordant |
| MLH1:c.2074T>C | p.(Ser692Pro) | Missense | PM2_supportin g, BP4, BS4 | Likely benign | Benign | Concordant |
| MSH2:c.1277-16T>C |  | Intronic | BP4, BS1 | Likely benign | Likely Benign | Concordant |
| MSH2:c.1748A>G | p.(Asn583Ser) | Missense | BP4, BS3 | Likely Benign | Likely Benign | Concordant |
| MSH2:c.913G>A | p.(Ala305Thr) | Missense | BP5, BS4_supportin g, BS3_supportin g, PP3_moderate, BS2 | Likely benign | Likely Benign | Concordant |
| MSH2:c.1563T>C | p.(Tyr521=) | Synonymous | BS1, BP4, BP7 | Likely benign | Likely Benign | Concordant |
| MLH1:c.2040C>T | p.(Cys680=) | Synonymous | PM2_supportin g, BP4, BP7 | Likely Benign | Likely Benign | Concordant |
| MLH1:c.277A>G | p.(Ser93Gly) | Missense | PM2_supportin g, BP4, BS3_supportin g | Likely benign | Likely Benign | Concordant |
| MSH6:c.3482CTG[1] | p.(Ala1162del) | Inframe deletion | PM2_supportin g, PS3, PP4 | Likely pathogenic | Likely pathogenic | Concordant |
| MLH1:c.1558+1G>A |  | Splice site | PM2_supportin g, PVS1 | Likely Pathogenic | Likely Pathogenic | Concordant |
| MSH2:c.2210+2T>C |  | Splice site | PM2_supportin g, PVS1 | Likely pathogenic | Likely pathogenic | Concordant |
| MSH2:c.792+1G>T |  | Splice site | PM2_supportin g, PVS1_strong, PP4 | Likely pathogenic | Likely Pathogenic | Concordant |
| MLH1:c.1646T>C | p.(Leu549Pro) | Missense | PP1_strong, PP3_moderate, PM2_supportin g | Likely pathogenic | Likely pathogenic | Concordant |
| PMS2:c.1144+1G>A |  | Splice site | PP4, PM2_supportin g, PVS1_strong | Likely pathogenic | Likely pathogenic | Concordant |
| MSH6:c.1296T>G | p.(Phe432Leu) | Missense | PS3, PP3, PM2_supportin g | Likely pathogenic | Likely Pathogenic | Concordant |
| PMS2:c.989-2A>G |  | Splice site | PVS1_strong, PP4_moderate, PM2_supporting | Likely pathogenic | Likely pathogenic | Concordant |
| MSH2:c.1807G>A | p.(Asp603Asn) | Missense | PM2_supporting, PP3, PP1, PP4_strong, PS3_moderate | Likely pathogenic | Pathogenic | Concordant |
| MSH6:c.1282A>G | p.(Lys428Glu) | Missense | PM2_supporting, PP3, PP4, PS3 | Likely pathogenic | Pathogenic | Concordant |
| MSH2:c.1862G>T | p.(Arg621Leu) | Missense | PM2_supporting, PP4_strong, PP3_moderate | Likely pathogenic | Pathogenic | Concordant |
| MSH6:c.1723G>T | p.(Asp575Tyr) | Missense | PM2_supporting, PP4_strong, PP3_moderate | Likely pathogenic | Pathogenic | Concordant |
| MLH1:c.350C>G | p.(Thr117Arg) | Missense | PM5, PM2_supporting, PP1_moderate, PP3_moderate, PP4_moderate | Likely pathogenic | Pathogenic | Concordant |
| MSH2:c.490G>T | p.(Gly164Trp) | Missense | PM5, PM2_supporting, PP3_moderate, PP4_moderate | Likely pathogenic | Pathogenic | Concordant |
| PMS2:c.241G>T | p.(Glu81Ter) | Nonsense | PVS1, PM2_supporting | Likely pathogenic | Pathogenic | Concordant |
| MLH1:c.790+1G>T |  | Splice site | PM2_supporting, PVS1, PP3 | Pathogenic | Likely Pathogenic | Concordant |
| MSH2:c.366+1G>C |  | Splice site | PM2_supporting, PVS1, PP1 | Pathogenic | Likely Pathogenic | Concordant |
| MSH2:c.929T>G | p.(Leu310Arg) | Missense | PM5, PM2_supporting, PP3_moderate, PP4_moderate, PP1_strong | Pathogenic | Likely Pathogenic | Concordant |
| MLH1:c.589-2A>C |  | Splice site | PVS1, PM2_supporting, PP4 | Pathogenic | Likely Pathogenic | Concordant |
| MSH2:c.1276G>A | p.(Gly426Arg) | Last base of exon | PM2_supporting, PP4, PVS1 | Pathogenic | Pathogenic | Concordant |
| MSH2:c.2075G>T | p.(Gly692Val) | Missense | PM2_supporting, PS3, PP3_moderate, PP4_strong | Pathogenic | Pathogenic | Concordant |
| PMS2:c.1571dup | p.(Gly525ArgfsTer17) | Frameshift | PM2_supporting, PVS1, PM3_supporting | Pathogenic | Pathogenic | Concordant |
| MSH2:c.1012G>A | p.(Gly338Arg) | Missense | PS3,<br>PM2_supportin<br>g, PP1,<br>PP3_moderate,<br>PP4_moderate | Pathogenic | Pathogenic | Concordant |
| MLH1:c.923A>C | p.(His308Pro) | Missense | PS3,<br>PP1_strong,<br>PM2_supportin<br>g, PP3_moderate | Pathogenic | Pathogenic | Concordant |
| MSH2:c.2459-12A>G |  | Intronic | PVS1,<br>PM2_supportin<br>g, PP4_strong | Pathogenic | Pathogenic | Concordant |
| PMS2:c.1753C>A | p.(Leu585Ile) | Missense | PM2_supportin<br>g, BP4 | Uncertain<br>significance | Uncertain<br>significance | Concordant |
| PMS2:c.614A>C | p.(Gln205Pro) | Missense | PM2_supportin<br>g, PM3, PP4,<br>PP3 | Uncertain<br>significance | Uncertain<br>significance | Concordant |
| MLH1:c.2041G>C | p.(Ala681Pro) | Missense | PM2_supportin<br>g, PP4 | Uncertain<br>significance | Uncertain<br>significance | Concordant |
| MLH1:c.2042C>T | p.(Ala681Val) | Missense | PP4_strong,<br>PM2_supportin<br>g | Uncertain<br>significance | Uncertain<br>significance | Concordant |
| MSH6:c.663A>C | p.(Glu221Asp) | Missense | BS1, BP4 | Likely benign | Uncertain<br>significance | Discordant |
| MSH2:c.2634+5G<br>>T |  | Intronic | PM2_supportin<br>g,<br>PP4_moderate,<br>PS1, PP3 | Likely<br>pathogenic | Uncertain<br>significance | Discordant |
| MLH1:c.1667G>A | p.(Ser556Asn) | Last base of<br>exon | PVS1_strong,<br>PP4_moderate,<br>PM2_supportin<br>g | Likely<br>pathogenic | Uncertain<br>significance | Discordant |
| MSH2:c.211G>C | p.(Gly71Arg) | Last base of<br>exon | PVS1,<br>PM2_supportin<br>g, PP4_strong | Pathogenic | Uncertain<br>significance | Discordant |
| MSH6:c.884A>G | p.(Lys295Arg) | Missense | BS4_supportin<br>g | Uncertain<br>significance | Likely Benign | Discordant |
| MLH1:c.1270G>A | p.(Ala424Thr) | Missense | PM2_supportin<br>g, BP4 | Uncertain<br>significance | Likely Benign | Discordant |
| MLH1:c.1963A>T | p.(Ile655Phe) | Missense | PM2_supportin<br>g, BP4 | Uncertain<br>significance | Likely Benign | Discordant |
| MLH1:c.1889T>A | p.(Ile630Asn) | Missense | PM2_supportin<br>g, PP1,<br>PP3_moderate,<br>PP4 | Uncertain<br>significance | Likely<br>Pathogenic | Discordant |

We compared the final ACMG/AMP-based classifications to the InSiGHT criteria classifications. It should be noted that the InSiGHT criteria included Bayesian methodology, which informed the VCEP on the number of tumors to count for PP4 and BP5 criteria, therefore general concordance was expected. Bayesian probabilities of pathogenicity that were calculated using tumor characteristics, segregation, and CIMRA functional data were compared to the ACMG/AMP classifications by converting the Bayesian probability using standard International Agency for Research on Cancer (IARC) probability categories for assigning B/LB (0.001/0.05) and P/LP (0.99/0.95).

The results show concordance of 40/48 (83%) variants between the ACMG/AMP and the InSiGHT criteria classifications. Reasons for discordance include four classifications that were achieved only with ACMG/AMP criteria evidence: PM5/PS1 (other variant at the same location classified as pathogenic), PVS1_Strong (last base of exon criteria), or BP4 (Missense variant with HCI combined MAPP+PolyPhen-2 prior probability). These evidence types were not used by the InSiGHT criteria. For three variants, the VCEP deemed that the InSiGHT classification using Bayesian probability was not appropriate due to insufficient clinical evidence, which also required changing the BP5 criteria to reflect this. Finally, one variant, *MLH1* c.1889T>A, reached 97% probability of pathogenicity based on tumor (PP4_Supporting), segregation data (PP1_Supporting), and *in silico* data (PP3_Moderate); however, this evidence combined with rare allele frequency (PM2_Supporting), Class 4 was not met using the ACMG/AMP combining rules. Overall, 47/48 (98%) of the ACMG/AMP-based classifications were concordant with InSiGHT outcomes or deemed by the VCEP to be more clinically appropriate.

Finally, the ACMG/AMP classifications were compared with clinical testing classifications on ClinVar. During which, 20/22 (91%) ClinVar variants were concordant with the ACMG/AMP classifications. An additional 23 variants on ClinVar had conflicting laboratory-submitted interpretations and 17 are now classified as P/LP or B/LB by this VCEP. The 3-star rating given to VCEP classifications will resolve the conflicting interpretations. One variant, *MLH1* c.2041G>C, was classified as LP on ClinVar but was a VUS using our ACMG/AMP criteria. The discrepancy was due, in part, to *in silico* analysis of protein structure showing a deleterious impact, reported by laboratories on ClinVar. However, the calibrated *in silico* prior probability used by this VCEP does not indicate an impact. Two variants, *MSH6* c.1723G>T and *MLH1* c.1646T>C, were reported on ClinVar as VUS from single submitters without any evidence provided, while the ACMG/AMP classification is LP for both. For these discordant variants, the VCEP 3-star classifications would take precedence over the existing laboratory-submitted classifications of LP.

## 4 Discussion

The VCEP approach of multidisciplinary experts and adoption of the ACMG/AMP criteria has been successful in achieving consensus and clinical validity for the complex task of variant interpretation (Ritter, 2019). The VCEP membership and classification process shares a strong overlap and similarity with the previous InSiGHT approach for variant classification. Adopting the ACMG/AMP criteria provides a solid foundation for organizing gene–disease-specific knowledge and experience and adds flexibility to use information that had not been formally calibrated for multifactorial likelihood analysis. In this work, we have implemented the ACMG/AMP-based criteria, superseding previous criteria for MMR genes. Our new ACMG/AMP criteria for LS-specific cancers were used to classify a pilot batch of 48 variants across a range of variant types, with diverse evidence to achieve pathogenic and benign classifications for 83% of variants. It is important to note that, owing to the selection of variants with available evidence for classification, it is not a representative sample of the whole variome for LS.

There are several important points to consider while using these criteria. The criteria apply to patients with LS-spectrum tumors, and specifically colorectal or endometrial cancer tumors for certain criteria. Although the CMMRD phenotype is a part of the criteria, variants should only be classified in the context of LS. We highlight that variants can be classified as B/LB for LS while causing the CMMRD phenotype in biallelic cases because of null/low penetrance in heterozygous carriers. Further development of the criteria is likely; therefore newer versions may provide different classifications of variants. Updated guidance and recommendations from ClinGen should also be considered alongside these criteria, as the ACMG/AMP criteria evolve due to advancements in variant classification, such as the recently proposed *in silico* analyses (Pejaver, 2022).

The advantages of the ACMG/AMP framework, as compared to our previous InSiGHT criteria, are evident. It is more flexible in combining disparate types of evidence than the InSiGHT criteria, which used predetermined combinations of evidence alongside a Bayesian multifactorial component. However, despite the ACMG/AMP criteria being compatible with Bayesian reasoning, in certain cases it may not reach a classification that the strictly Bayesian multifactorial likelihood calculation approach would provide. This may be a possible avenue for fine-tuning of the criteria. Nevertheless, the new ACMG/AMP criteria for the MMR genes, with set minimum thresholds for evidence (e.g., BA1, BS1 allele frequency thresholds, *in silico* odds, tumor characteristics, functional assay odds, and co-segregation odds), varied evidence types, and logical combining rules, will fill a valuable role in the classification process for clinical genetics. A purely quantitative approach might have less usability in routine clinical diagnostics due to inherent complexity or impracticality. This is evident in the global uptake of the ACMG/AMP criteria and the sharing of classification criteria between different VCEPs for improving the criteria over time. Overall, these new criteria complement the MMR-specific Bayesian multifactorial likelihood methods and supersede the previous InSiGHT classification criteria for MMR genes, providing capacity for wider clinical application. This VCEP supports expert specifications to the original ACMG/AMP criteria, owing to elaborate gene-specific criteria for frequently analyzed genes, such as MMR, to improve clinical care.

## 5 Conclusion

The ACMG/AMP criteria offer a flexible and standardized system for MMR variant classification, with gene–disease-specific knowledge to enhance the criteria. By adopting existing recommendations and incorporating efforts from functional assays, computational analyses, and clinical domains, a coherent and practical set of criteria for variant classification was achieved for MMR genes, using the ACMG/AMP framework. These MMR-specific ACMG/AMP criteria set a foundation, with the ability to include additional *in silico* predictors, functional assays, and clinical evidence, as science advances over time. The InSiGHT ClinGen VCEP will employ these criteria for ongoing MMR variant classifications and will seek to update and enhance the criteria in alignment with ClinGen and other VCEP groups.

## 6 Data availability

Variant classifications are published in ClinVar (https://www.ncbi.nlm.nih.gov/clinvar/) and curated evidence is available on the ClinGen Evidence Repository (https://erepo.clinicalgenome.org/evrepo/).

## 7 Acknowledgments

JPP was supported by the Alan Watt and Chris Geyer Oncology Fellowship through The Royal Melbourne Hospital Foundation and NIH PA-18-591 Administrative Supplements to Promote Data Sharing in Cancer Epidemiology Studies. ABS was supported by NHMRC Funding (APP1104808). Grant funding for MKC: Cancer Council NSW, RG19-1. In Spain, this study has been partially funded by Ministerio de Ciencia e Innovación, which is part of Agencia Estatal de Investigación (AEI), through the Retos Investigación grant, number PID2019-111254RB-I00, and La Marató de TV3 (202028-30). We also thank CERCA Programme / Generalitat de Catalunya for institutional support. MG was supported by Italian Ministry of Health RC 2022. KA and GY was supported by Japan Agency Medical Research and Development (AMED) under grant JP 18kk0205004, JSPS KAKENHI Grant Number JP18K07339 and JP22K07266. We would like to thank Ms. Lubaina Koti for editing the manuscript for language, structure, and accuracy. We acknowledge Marissa Rose for analysis of an early draft of the criteria.

## 8 Conflict of Interests

Becky Milewski previously had a role with the pilot variant classification efforts in 2020/2021. She was previously an employee of GeneDx, LLC.

Lauren Currie is an employee of GeneDx, LLC

Ester Borras is an employee and shareholder of Invitae Corporation.

John Paul Plazzer has previously consulted as curator for an external diagnostic testing company.

## List of abbreviations

ACMG: American College of Medical Genetics and Genomics
AMP: Association for Molecular Pathology
B: Benign
BA: Benign Stand-Alone
BS: Benign Strong
BP: Benign Supporting
ClinGen: Clinical Genome Resource
CMMRD: constitutional mismatch repair deficiency
GI: gastrointestinal
gnomAD: Genome Aggregation Database
FDA: Food and Drug Administration (United States)
HCI: Huntsman Cancer Institute
HGVS: Human Genome Variation Society
IARC: International Agency for Research on Cancer
IHC: immunohistochemistry
InSiGHT: International Society for Gastrointestinal Hereditary Tumours
IVS: intervening sequence (intron)
LB: likely benign
LP: likely pathogenic
LOVD: Leiden Open Variation Database
LS: Lynch syndrome
MANE: matched annotation from National Center for Biotechnology Information and European Molecular Biology Laboratory and European Bioinformatics Institute
MMR: mismatch repair (genes)
MSI: microsatellite instability
NGS: next generation sequencing
NMD: nonsense–mediated mRNA decay
P: Pathogenic
PVS: Pathogenic Very Strong
PS: Pathogenic Strong
PM: Pathogenic Moderate
PP: Pathogenic Supporting
SVI WG: Sequence Variant Interpretation Working Group
VCEP: variant curation expert panel
VUS: variant of uncertain significance

---

Supplementary Table 1. Validation of functional assays (See supplementary excel file).

## APPENDIX

### Important Notes

*PMS2* NGS results need confirmation by other orthogonal assays as well as functional assessment (e.g. Long-Range or cDNA), if variants are located in the *PMS2CL* pseudogene homologous regions (exons 11-15).

Gene-specific penetrance estimates are available at http://lscarisk.org/

### Justification for last exon PVS1 boundaries

Nonsense/frameshift variant introducing Premature Termination Codon (PTC):

1. ≤ codon 753 in MLH1 using location of known pathogenic variant *MLH1* c.2252_2253del
2. ≤ codon 891 in MSH2 using location of known pathogenic variant *MSH2* c.2662del
3. ≤ codon 1341 in MSH6 using location of known pathogenic variant *MSH6* c.3984_3987dup
4. ≤ codon 798 in PMS2 using ≥50 nucleotide NMD-rule.

### Protein Expression and consistency with variant location

IHC evidence should be consistent with the variant gene and the protein that is tested and take into account the MutSα and MutLα heterodimers: MLH1 and PMS2 loss is consistent with an *MLH1* pathogenic variant, MSH2 and MSH6 loss is consistent with an *MSH2* pathogenic variant, MSH6 loss is consistent with an *MSH6* pathogenic variant, and PMS2 loss is consistent with a *PMS2* pathogenic variant.

## Notes

### Author Declarations

Data is available in published literature (Pubmed IDs are provided in the databases listed here) or online open-access databases including: ClinVar https://www.ncbi.nlm.nih.gov/clinvar/, LOVD https://databases.lovd.nl/shared/, and InSiGHT classification websites: https://www.insight-database.org/classifications/mmr_integrative_eval.html and https://www.insight-database.org/classifications/ Curated evidence will be available on https://erepo.clinicalgenome.org/evrepo/

## References

André, T., 2020. Pembrolizumab in Microsatellite-Instability–High Advanced Colorectal Cancer. New England Journal of Medicine, pp. 2207–2218.

Aronson, M., 2022. Diagnostic criteria for constitutional mismatch repair deficiency (CMMRD): recommendations from the international consensus working group. Medical Genetics, pp. 318–327.

Belman, S., 2020. Considerations in assessing germline variant pathogenicity using cosegregation analysis. Genetics in Medicine, 22(12), pp. 2052–2059.

Biesecker, L. a. H. S., 2018. The ACMG/AMP reputable source criteria for the interpretation of sequence variants. Genetics in Medicine, 20(12), pp. 1687–1688.

Biller, L., 2022. Lynch Syndrome-Associated Cancers Beyond Colorectal Cancer. Gastrointestinal Endoscopy Clinics of North America, pp. 75–93.

Brnich, S., 2020. Recommendations for application of the functional evidence PS3/BS3 criterion using the ACMG/AMP sequence variant interpretation framework. Genome medicine, 12(1), pp. 1–12.

Canson, D., 2022. The splicing effect of variants at branchpoint elements in cancer genes. Genetics in Medicine, 24(2), pp. 398–409.

Cyr, J., 2012. The predicted truncation from a cancer-associated variant of the MSH2 initiation codon alters activity of the MSH2-MSH6 mismatch repair complex. Molecular carcinogenesis, 51(8), pp. 647–658.

Dominguez-Valentin, M., 2020. Cancer risks by gene, age, and gender in 6350 carriers of pathogenic mismatch repair variants: findings from the Prospective Lynch Syndrome Database. Genetics in Medicine, 22(1), pp. 15–25.

Dominguez-Valentin, M., 2021. No difference in penetrance between truncating and missense/aberrant splicing pathogenic variants in MLH1 and MSH2: a prospective lynch syndrome database study. Journal of clinical medicine, 10(13), p. 2856.

Dominguez-Valentin, M., 2023. Mortality by age, gene and gender in carriers of pathogenic mismatch repair gene variants receiving surveillance for early cancer diagnosis and treatment: a report from the prospective Lynch syndrome database. EClinicalMedicine.

Drost, M., 2019. A functional assay–based procedure to classify mismatch repair gene variants in Lynch syndrome. Genetics in Medicine, 21(7), pp. 1486–1496.

Drost, M., 2020. Two integrated and highly predictive functional analysis-based procedures for the classification of MSH6 variants in Lynch syndrome. Genetics in Medicine, 22(5), pp. 847–856.

Fokkema, I., 2021. The LOVD3 platform: efficient genome-wide sharing of genetic variants. European Journal of Human Genetics, 29(12), pp. 1796–1803.

Georgeson, P., 2019. Tumor mutational signatures in sebaceous skin lesions from individuals with Lynch syndrome. Molecular Genetics & Genomic Medicine, p. e00781.

Jia, X., 2021. Massively parallel functional testing of MSH2 missense variants conferring Lynch syndrome risk. The American Journal of Human Genetics, 108(1), pp. 163–175.

Kahn, R., 2019. Universal endometrial cancer tumor typing: How much has immunohistochemistry, microsatellite instability, and MLH1 methylation improved the diagnosis of Lynch syndrome across the population?. Cancer, 125(18), pp. 3172–3183.

Karczewski, K. J. F. L. C. T. G. C. B. B. A. J. W. Q. .. &. M. D. G., 2020. The mutational constraint spectrum quantified from variation in 141,456 humans.. Nature, 581(7809), pp. 434–443.

Latham, A., 2019. Microsatellite instability is associated with the presence of Lynch syndrome pan-cancer. Journal of clinical oncology, 37(4), p. 286.

Lewis, B., 2003. Evidence for the widespread coupling of alternative splicing and nonsense-mediated mRNA decay in humans. Proceedings of the National Academy of Sciences, 100(1), pp. 189–192.

Li, S., 2020. Tumour characteristics provide evidence for germline mismatch repair missense variant pathogenicity. Journal of Medical Genetics, 57(1), pp. 62–69.

Nugroho, P., 2022. Risk of cancer in individuals with Lynch-like syndrome and their families: a systematic review. Journal of Cancer Research and Clinical Oncology, pp. 1–22.

Pejaver, V., 2022. Calibration of computational tools for missense variant pathogenicity classification and ClinGen recommendations for PP3/BP4 criteria. The American Journal of Human Genetics.

Plazzer, J., 2013. The InSiGHT database: utilizing 100 years of insights into Lynch syndrome. Familial cancer, pp. 175–180.

Preston, C., 2022. ClinGen Variant Curation Interface: a variant classification platform for the application of evidence criteria from ACMG/AMP guidelines. Genome Medicine, 14(1), pp. 1–12.

Rath A, R. A. R. K. G. R. H. J. C. M. T. S. G. J. H. C., 2022. A calibrated cell-based functional assay to aide classification of MLH1 DNA mismatch repair gene variants. Human Mutation, 43(12), pp. 2295–2307.

Rayner, E., 2022. Predictive functional assay-based classification of PMS2 variants in Lynch syndrome. Human Mutation, 43(9), pp. 1249–1258.

Reese, M. G. E. F. H. K. D. &. H. D., 1997. Improved splice site detection in Genie. Proceedings of the first annual international conference on Computational molecular biology, pp. 232–240.

Rehm, H., 2015. ClinGen—the clinical genome resource. New England Journal of Medicine, 372(23), pp. 2235–2242.

Richardson, M., 2019. DNA breakpoint assay reveals a majority of gross duplications occur in tandem reducing VUS classifications in breast cancer predisposition genes. Genetics in Medicine, 21(3), pp. 683–693.

Richards, S., 2015. Standards and guidelines for the interpretation of sequence variants: a joint consensus recommendation of the American College of Medical Genetics and Genomics and the Association for Molecular Pathology. Genetics in medicine, pp. 405–423.

Riepe, T., 2021. Benchmarking deep learning splice prediction tools using functional splice assays. Human Mutation, 42(7), pp. 799–810.

Ritter, D., 2019. A case for expert curation: an overview of cancer curation in the clinical genome resource (ClinGen). Molecular Case Studies, 5(5), p. a004739.

Rivera-Muñoz, E., 2018. ClinGen Variant Curation Expert Panel experiences and standardized processes for disease and gene-level specification of the ACMG/AMP guidelines for sequence variant interpretation. Human Mutation, pp. 1614–1622.

Scott, A., 2022. Saturation-scale functional evidence supports clinical variant interpretation in Lynch syndrome. Genome Biology, 23(1), pp. 1–16.

Strande, N., 2018. Navigating the nuances of clinical sequence variant interpretation in Mendelian disease.. Genetics in Medicine, 20(9), pp. 918–926.

Tavtigian, S., 2018. Modeling the ACMG/AMP variant classification guidelines as a Bayesian classification framework. Genetics in Medicine, 20(9), pp. 1054–1060.

Tayoun, A., 2018. Recommendations for interpreting the loss of function PVS1 ACMG/AMP variant criterion. Human mutation, 39(11), pp. 1517–1524.

Thompson, B., 2013. A multifactorial likelihood model for MMR gene variant classification incorporating probabilities based on sequence bioinformatics and tumor characteristics: a report from the Colon Cancer Family Registry. Human Mutation, 34(1), pp. 200–209.

Thompson, B., 2014. Application of a 5-tiered scheme for standardized classification of 2,360 unique mismatch repair gene variants in the InSiGHT locus-specific database. Nature genetics, pp. 107–115.

Thompson, B., 2020. Contribution of mRNA splicing to mismatch repair gene sequence variant interpretation. *Frontiers in genetics*, Volume 11, p. 798.

Tian, Y., 2019. REVEL and BayesDel outperform other in silico meta-predictors for clinical variant classification. Scientific reports, 9(1), pp. 1–6.

Tiwari, A., 2016. Lynch syndrome in the 21st century: clinical perspectives. QJM: An International Journal of Medicine, pp. 151–158.

Whiffin, N., 2017. Using high-resolution variant frequencies to empower clinical genome interpretation. Genetics in Medicine, 19(10), pp. 1151–1158.

Yeo, G. &. B. C. B., 2004. Maximum entropy modeling of short sequence motifs with applications to RNA splicing signals. J Comput Biol, 11(2-3), pp. 377–94.

Zhang, Y., 2018. A Novel MLH1 Initiation Codon Mutation (c.3G>T) in a Large Chinese Lynch Syndrome Family with Different Onset Age and mRNA Expression Level. BioMed Research International.

